# Execution of intervention matters more than strategy: A lesson from the spatiotemporal assessment of COVID-19 clusters in Nepal

**DOI:** 10.1101/2020.11.07.20227520

**Authors:** Bipin Kumar Acharya, Laxman Khanal, Alnwisi Sameh Mansoor Mahyoub, Zengliang Ruan, Yin Yang, Samir Kumar Adhikari, Shreejana Pandit, Basanta Kumar Neupane, Binod Kumar Paudel, Hualiang Lin

**Author notes:** **Correspondence:** Huliang Lin. Two authors contributed to the manuscript equally (Bipin Kumar Acharya, Laxman Khanal).

## Abstract

The novel coronavirus disease 2019 (COVID-19) has been the biggest public health problem of the present world. As the number of people suffering from the pandemic is rising, it is likely to claim more life and worsen the global health and economy. Nepal, one of the developing countries in the south Asia has been strongly influenced by the pandemic and struggling to contain it with multiple interventions, however, spatiotemporal dynamics of the epidemic and its linkage with various intervention strategies has not been studied yet. Here, we employed the prospective spatial-temporal analysis with SaTScan assessing dynamics of the COVID-19 cases from 23 January to 31 August 2020 at district level in Nepal. The results revealed that COVID-19 dynamics in the early stage of transmission was slower and confined in certain districts. However, from the third week of April, transmission spread rapidly across districts of Province No. 2 and Sudoorpaschim Province, primarily introduced by Nepalese citizens returning from India. Despite nationwide lockdown, nine statistically significant active and emerging clusters were detected between 23 January and 21 July 2020, whereas ten emerging clusters were observed for extended period to 31 August. The population density and population inflow from India crossing the sealed border had significant effects on the elevated risk of the epidemic. The capital city Kathmandu has become the highest-risk active cluster since August when travel restriction has been suspended. Movement restriction appears to be the most effective non-pharmaceutical intervention against the COVID-19 for resource-scarce countries with limited health care facilities. Our findings could be valuable to the health authorities within Nepal and beyond to better allocate resources and improve interventions on the pandemic for containing it efficiently.

## 1. Introduction

The coronavirus disease 2019 (COVID-19) outbreak has been considered a Public Health Emergency of International Concern on 30 January 2020 and declared a global pandemic on 11 March 2020 by the World Health Organization (WHO). This viral disease caused by the severe acute respiratory syndrome coronavirus 2 (SARS-CoV-2) has infected 39,596,858 people and killed 1107374 lives as of 18 October 2020 (WHO, 2020). COVID-19 is a highly contagious disease with an initial estimated average basic reproductive rate (R_0_) of 3.28 (Liu et al., 2020) that has been substantially reduced by the multiple intervention approaches (You et al., 2020). The disease has been described to have higher severity to the old-age and immunosuppressed people such as suffering from cardiac and pulmonary disorders (Zheng et al., 2020). Despite the constant efforts of scientists across the globe, we still lack the well-tested and approved vaccines and drugs against the disease.

Analogous to many countries in the world, Nepal has hugely suffered from the COVID-19 pandemic (Panthee et al., 2020). The first case of the disease was reported on 23 January 2020 in a China-returned 32 years old male (Pun et al., 2020; Shrestha et al., 2020). After the lag period of two months, the second positive case was reported on 23 March 2020 (Piryani et al., 2020). Subsequently the number of the disease victims gradually increased and almost all the cases in the earlier days were among the people returned from Europe, Middle-East countries and others. Despite being neighbored to the China also, the number of SARS-CoV-2 positive cases in Nepal surged up when the Nepalese workers from its southern neighbor India returned home via open border between the two countries. By the date of 18 October 2020, there have been 132246 cases and 739 deaths across the country (MoHP, 2020). The geographical distribution of the COVID-19 cases within the Nepal territory is not uniform. Communities associated with the poverty in densely populated growing cities are estimated more vulnerable to the infection (Khanal et al., 2020). Additionally, disease intervention efforts are not equal among the local administrative units of Nepal and so is the pattern of spread of the COVID-19.

The magnitude and timing of the interventions matter for the mitigation of the outbreak (Dehning et al., 2020). When the second COVID-19 case was recorded in Nepal and number of cases was also rising in India, the Government of Nepal closed all international flights and borders on 23 March 2020 (Sapkota et al., 2020). The very next day, a nation-wide lockdown was further enforced that continued till 21 July 2020. Besides the diagnosis, isolation and treatment of the COVID-19 patients, Government of Nepal employed multiple public health measures such as border closure, lockdown, social distancing, and personal hygiene which aided Nepal in avoiding the spread of the novel coronavirus during the initial days (Basnet et al., 2020; Dhakal and Karki, 2020). Physical distancing measures, such as closure of schools and colleges, retail businesses, and restaurants, cancellation of public events, as well as constraints on individual movements and social interactions, are now in place in many countries with the aim of reducing transmission of SARS-CoV-2 (Cowling et al., 2020; Davies et al., 2020; Yang et al., 2020). Among other measures, travel restrictions, physical distancing, home quarantine, centralized quarantine, compulsion on mask wearing in public places, universal symptom survey, implementation of testing, isolation, and contact tracing probably slowed the transmission dynamics significantly (Davies et al., 2020; Fang et al., 2020; Pan et al., 2020). Despite of continuous lockdown enforced for 120 days, the outbreak of the COVID-19 is reemerging and the efficacy of such interventions have never been assessed in Nepal.

Pharmaceutical interventions alone are not enough to contain the COVID-19, hence, countries augmented them with non-pharmaceutical approaches. However, how different combinations of interventions, timings, and extents have yielded desired outcomes to curb the disease transmission remains unclear (Cowling et al., 2020; Davies et al., 2020; Fang et al., 2020; MacIntyre and Wang 2020; Pan et al., 2020; Zhang et al., 2020). The level of vulnerability to the COVID-19 differed among the communities based on the demographic, socioeconomic, accessibility to the health facilities, prevalence of the pulmonary and cardiac disorders, etc. (Khanal et al., 2020). The complex spatial and temporal epidemiology of COVID-19 due to rapid changes in human population dynamics and its demographic and environmental drivers challenges in its efficient control (Alkhamis et al., 2020). One of the important drivers of the spreading of infectious diseases is the human movement, tracking of which is using data sources such as public transportation (bus, train, and flight), social-media data, and mobile-phone data could be critical for the prediction of virus transmission, the identification of risk area, and decisions about control measure (Zhou et al., 2020). Therefore, it is important to analyze the spatiotemporal pattern of the COVID-19 outbreak in the light of human dynamics and non-pharmaceutical interventions.

The use of spatiotemporal analytical tools for rapid risk-based surveillance can offer valuable near-real-time insights into the severity of pandemic spread as well as the effectiveness of intervention measures and can aid decision making, planning and community action (Franch-Pardo et al., 2020). Such analyses have been proven to be effective to identify the extent and impact of the pandemic and formulate the intervention strategies in many countries including Bangladesh (Masrur et al., 2020), China (Kang et al., 2020; Xie et al., 2020), Italy (Gatto et al., 2020; Martellucci et al., 2020), Spain (Santamaría and Hortal 2021), USA (Cordes and Castro 2020; Mollalo et al., 2020; Sun et al. 2020), etc. Additionally, findings of such analyses could be decisive for early identification of high-risk groups for disease transmission and efficient deployment of resources for developing economies like Nepal. The spatial scan statistic (SaTScan) is a widely used method for geographical disease surveillance that detects and determines statistical significance of geographical cluster without having to prespecify the cluster size or location. It applies a moving circular window on the map, centered on each of many possible grid points positioned throughout the study region and creates many thousands of distinct geographical circles, each being the possible candidate cluster. The retrospective analysis on the SaTScan can identify all past and current significant clustering events throughout the study period (Kulldorff, 1997).

The epidemic crisis management demands estimation of the actual effects of interventions taken not only to make rapid adjustments but also to adapt short-term forecasts (Dehning et al., 2020). Nepal offers an opportunity to assess the impact of non-pharmaceutical interventions on COVID-19 that could be rolled out in resource-limited settings in other countries. The main objective of this study was to assess spatiotemporal dynamics of COVID-19 on the context of various restrictions imposed as preventive measures to contain the disease transmission. We explored active and emerging disease clusters using the prospective space–time scanning (Desjardins et al., 2020; Masrur et al., 2020) for two time periods, 23 January - 21 July, and 23 January - 31 August 2020 taking the cutoff date of 21 July. In addition, we investigated biweekly space-time propagation of transmission for locating risk and newly emerged clusters along the timeline accounting two weeks incubation period of the SARS-CoV-2.

## 2. Methods

### 2.1 Data Source

This study was conducted covering the entire 77 districts of Nepal (Fig. 1) using three different datasets. COVID-19 cases reported to the Ministry of Health and Population (MoHP), Government of Nepal was the first of its kind. This dataset contains daily COVID-19 positive cases, death and recovery aggregated at district. The COVID-19 were tested using the RT-PCR in various lab distributed across the country. We extracted reported positive case and joined them with district shapefile collected from Department of Survey, Government of Nepal. In addition, we obtained gridded population dataset in 100-meter spatial resolution for the year of 2020 from the worldpop geoportal (https://www.worldpop.org/). We summarized it for each district using the zonal statistics tool of ARC GIS which was used later as a base population to assess underlying risk to COVID-19 in the district.

**Figure 1.**
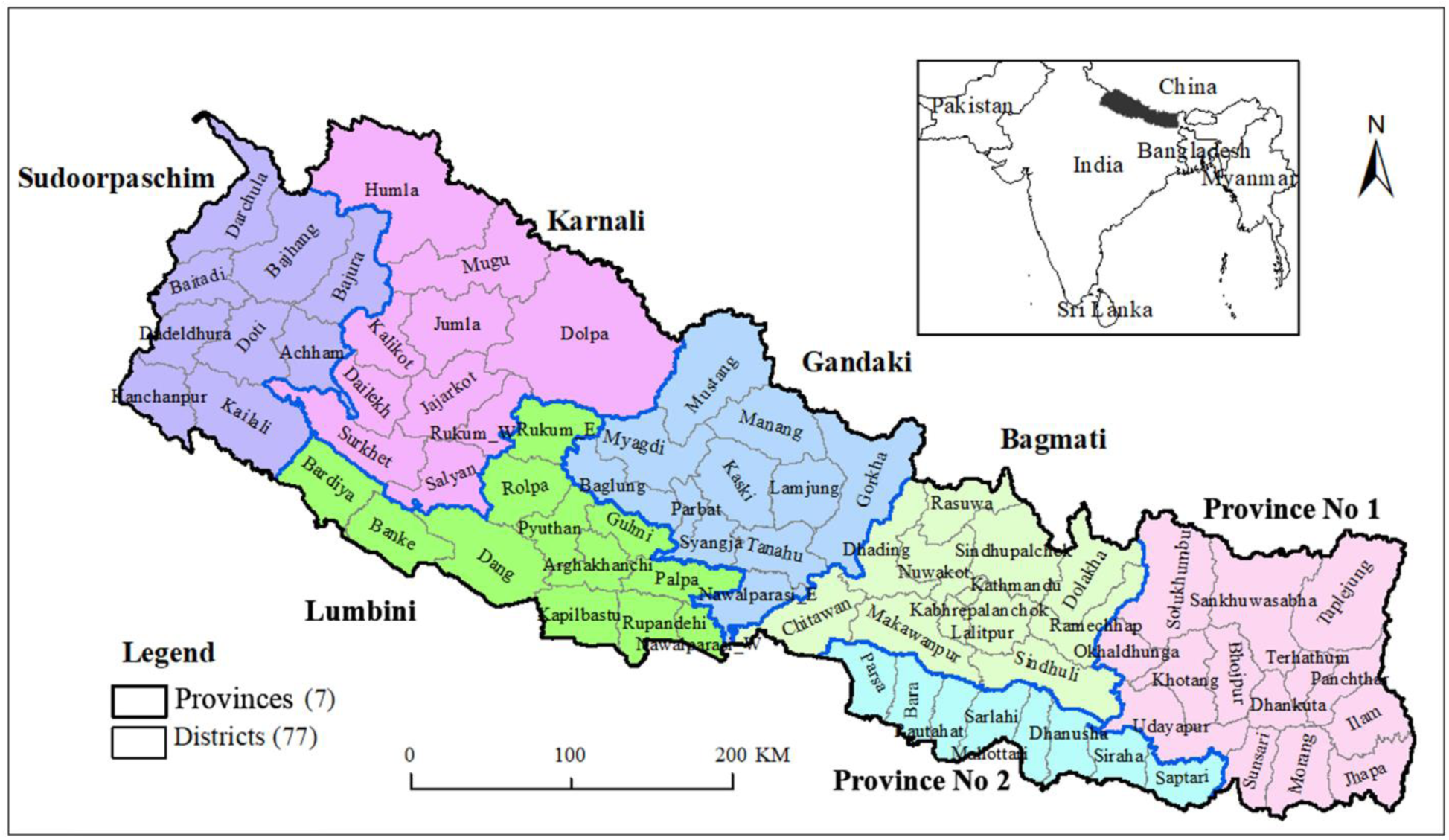
Location map of Nepal showing the seven provinces and 77 districts.

### 2.2 Data Analysis

We used geospatial analytics to characterize spatiotemporal dynamics of COVID-19 in Nepal. We divided study period in two parts based on the national level lockdown employed by the federal government of Nepal. The country was shut down from 23 March 2020, which was lifted five months later in 21 July 2020. We visualized the temporal dynamics using epidemic curve and several restrictions enforced in different spatiotemporal scales. The spatial distribution of cumulative cases of reported COVID-19 and incidence rate before and after the cutoff date of 21 July 2020 is presented through choropleth mapping technique.

To quantify spatiotemporal dynamics of the epidemics, we used the SatScan approach (Kulldorff, 1997) using the SaTScan version 9.6 (Kulldorff, 2018). The SaTScan statistics has been used widely to identify significant spatial/ temporal and spatiotemporal disease clusters including COVID-19 in different region of the world (Acharya et al., 2016; Desjardins et al., 2020; Masrur et al., 2020). The SaTScan scans across time and/or space using moving window to identify possible clusters by comparing the number of observed cases and expected cases assuming random distribution inside the window at each location. Scanning window is a time interval for purely temporal scan, a circle or ellipse in spatial scan and a cylinder in space-time scan where base of a cylinder represents space dimension and height represents the temporal dimension (Kulldorff, 2001; Kulldorff, 2018). The null hypothesis states that there is no difference in risk between inside and outside of the circle or cylinder while alternative hypothesis states the number of observed cases exceeds the number of expected cases derived from null models with elevated risk within the circle/cylinder. The expected number (*μ*) under the null hypothesis H_0_ is derived as follows (Desjardins et al., 2020):

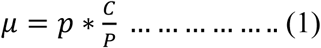

where, *p* is the population in a geographic area, C and P are the total number of reported cases and the total estimated population, respectively.

The SaTScan also identifies secondary clusters in addition to the most likely cluster for spatial and spatiotemporal scan, and orders them according to their likelihood ratio test. Equation (2) was used for calculating maximum likelihood ratio that identified scanning windows with elevated risk (Kulldorff, 2001).

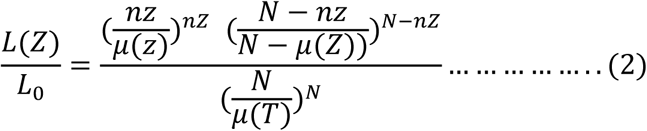

where, L(Z)= Log likelihood function for scanning window; L_0_ is the likelihood function for null hypothesis; nZ is the number of reported cases within the window; µ(Z) is the number of expected cases within the window Z; N is the total number of observed cases for the entire study areas across all time periods; and µ(T) is the total number of expected cases in the study area across all time periods.

Due to the assumption of uniform relative risk (RR) across a cluster, multiple geographic units can belong to significant space-time clusters. To avoid that assumption, relative risk for each spatial unit that belongs to a cluster is computed. The relative risk is for each location belonging to a cluster is calculated as (Liu et al., 2018; Hohl et al., 2020):

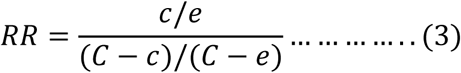

where, ‘c’ is the total number reported cases, ‘e’ is the total number of expected cases, and ‘C’ is the total number of observed cases in the entire study area.

In this study, we chose prospective space time analysis to detect emerging or active space-time clusters that are still occurring at the end of the study period based on the discrete Poisson model (Kulldorff, 1997, 2001). We chose discrete Poisson probability model to account heterogeneous distribution of COVID-19 transmission across the space and time (Kim and Castro 2020; Masrur et al., 2020). The space-time scan statistic employs moving cylinders for potential space-time clusters of COVID-19 cases. We performed this analysis on daily reported cases of COVID-19 aggregated on 77 districts. As we were interested to locate elevated risk zones to the COVID-19, high rate was chosen for further analysis. We set the upper bounds to have a maximum spatial and temporal scanning window size of 10% of the population at-risk to avoid extremely large clusters; and 50% of the study period, respectively. We utilized Monte Carlo testing with 9999 replications to assess the statistical significance of space-time clusters with default P of 0.05.

To understand the space-time propagation of the transmission we computed difference of relative risk between two-study periods and also detected emerging clusters using with shorter temporal scan through biweekly cumulative prospective scanning approach accounting two weeks incubation period (Desjardins et al., 2020) for locating the risk and newly emerged high-risk areas along the timeline.

## 3. Results

### 3.1 General overview of the COVID-19 in Nepal

A total 39460 cases of COVID-19 were reported in Nepal as of 31 August 2020 out of 693,472 tests done by RT-PCR. Among the total infected people, 30,881 (78.25%) were males and 8579 (21.74%) were females. The fatality rate and cured rate in Nepal due to COVID-19 were 0.6% and 54.3%, respectively.

The temporal dynamics of the epidemic is presented in the Fig. 2. The epidemic curve started ascend only after third week of April, which was almost four months later the detection of the first case in 23 January 2020. However, sporadically COVID-19 cases were detected from different districts despite nationwide lockdown started on 24 March. From the third week of the May the epidemic curve started to rise abruptly and the trend continued until the June last. In this period, significantly higher number of migrant workers returned home from India. Once the number of returns from India decreased slowly the positive case also shrunk rapidly. However, the number of COVID-19 cases increased substantially again after lifting the nationwide lockdown in 21^st^ July 2020. The rising trend continued in the later part of July and entire August despite the local level restrictions enforced in different districts.

**Figure 2.**
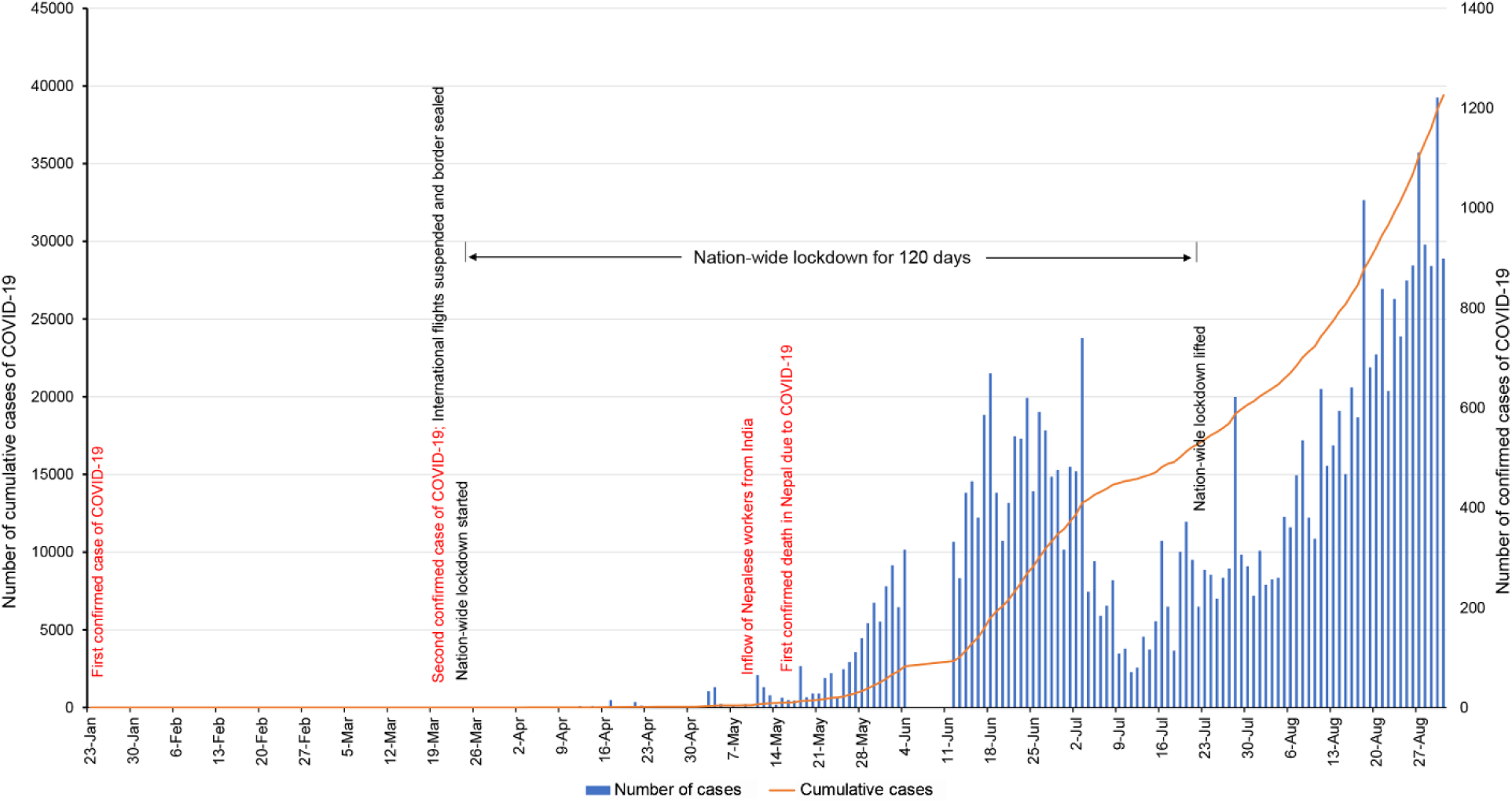
Epidictic curve of COVID-19 cases in Nepal reported. The primary axis (left) is cumulative count of reported cases and secondary axis (right) is daily reported count

Spatial distribution of cumulative cases and district level incidence rate of COVID-19 reported before and after the cutoff date is presented in the Figures 3a and 3b, respectively. By July 21, the epidemic was more intense in several western districts such as Dailekh, Doti, Achham and Bajura and low lying Tarai districts bordering with India including Rautahat, Kailali, Mahotari and Sarlahi, although it was already spread across the country. Spatial pattern of the incidence rate was slightly different than the patterns of total cumulative cases which determined by the population distribution. Bajura, Doti, Achham, Dailekh were districts with higher incidence. Higher incidence rates were also reported from Palpa, Parbat and Arghaghkanchi districts of Gandaki province. By August 31, the epidemic had become more intense across the country (Figure 3b). The highest number of cases were reported from Kathmandu followed by Parsa, Sarlahi, Rautahat while the least cases were reported from Mustang, Manang and Humla, respectively. In the same period, the highest incidence was observed in Doti, followed by Bajura and Dailekh, where the incidence was above 30/1000.

**Figure 3.**
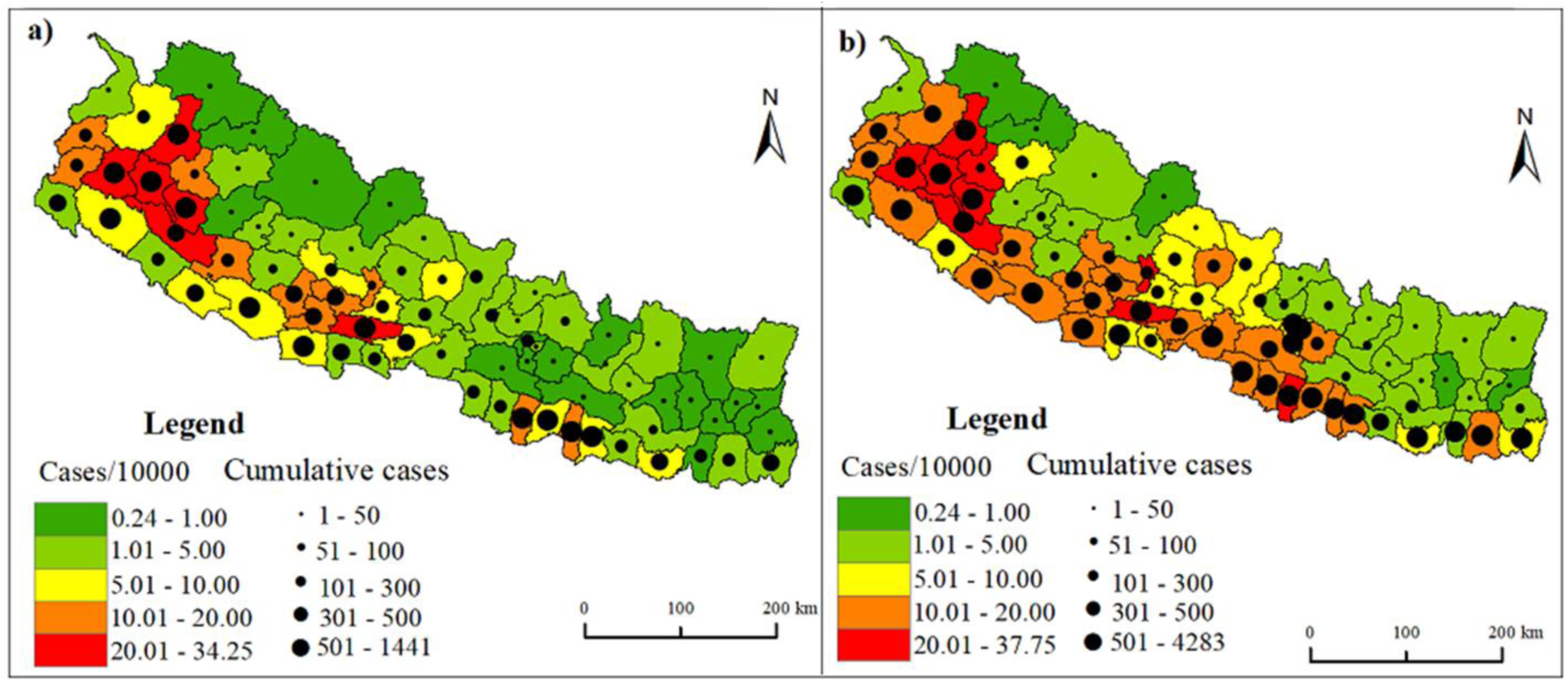
Spatial distribution of cumulative number and rate of incidence/10000 of COVID-19 cases from a) 23 January – 21 July and b) 23 January – 31 August 2020

### 3.2 Emerging district level clusters: 23 January-21 July 2020

Nine statistically significant emerging space-time clusters of COVID-19 were detected at district level between January 23 and July 21, 2020 in Nepal. Table 1 provides the characteristics of these clusters with varying size, relative risk and onset time and duration. The most likely cluster i.e. cluster 1 and other secondary clusters; 3, 4 and 5 emerged from June 12 while cluster 6, 7 and 8 lately emerged almost at the end of study period. The relative risk of these clusters also varied significantly. For example, the RR of cluster 1 (most likely cluster) was 16.95 while those of the cluster 2 and 3 were 9.87 and 9.81, respectively. Cluster 5, 8 and 9 were low risk clusters with RR less than 3.00.

**Table 1.** District level emerging space-time clusters of COVID-19 from 23 January to 21 July 2020 in Nepal (RR: relative risk). All results are statistically significant at P<0.001.

| Cluster | Radius | Start Date | End Date | # Districts | Observed | Expected | RR |
| --- | --- | --- | --- | --- | --- | --- | --- |
| 1 | 82.89 | 2020/6/12 | 2020/7/21 | 11 | 4574 | 361.58 | 16.95 |
| 2 | 145.18 | 2020/6/14 | 2020/7/21 | 19 | 2884 | 344.79 | 9.87 |
| 3 | 53.05 | 2020/6/12 | 2020/7/21 | 5 | 2843 | 341.13 | 9.81 |
| 4 | 0.00 | 2020/6/12 | 2020/7/21 | 1 | 307 | 67.52 | 4.61 |
| 5 | 44.18 | 2020/6/12 | 2020/7/21 | 3 | 386 | 222.55 | 1.75 |
| 6 | 27.29 | 2020/7/21 | 2020/7/21 | 3 | 36 | 8.08 | 4.46 |
| 7 | 51.75 | 2020/7/16 | 2020/7/21 | 2 | 22 | 2.97 | 7.40 |
| 8 | 51.33 | 2020/7/20 | 2020/7/21 | 4 | 81 | 36.07 | 2.25 |
| 9 | 0.00 | 2020/6/23 | 2020/7/21 | 1 | 50 | 20.77 | 2.41 |

Figure 4a shows the locations and spatial patterns of the nine emerging space-time clusters of COVID-19 at the district level in Nepal between January 23 and July 23, 2020. Cluster 1 contains 11 districts of Karnali and Sudoorpaschim province. Cluster 2, the first secondary cluster, is the largest cluster with 145 km radius and covers 19 districts of western Nepal. Cluster 3, 5 and 6 were smaller compared to the first two clusters with radius 53, 44 and 27 km and number of districts inside the clusters were 5, 3 and 3, respectively. Cluster 4 and 9 were single district cluster of Saptari and Sindhupalchok, correspondingly. There were 28 out of 77 districts outside these 9-emerging clusters having RR=0; at the time of this analysis, they were non emerging COVID-19 risk districts.

**Figure 4.**
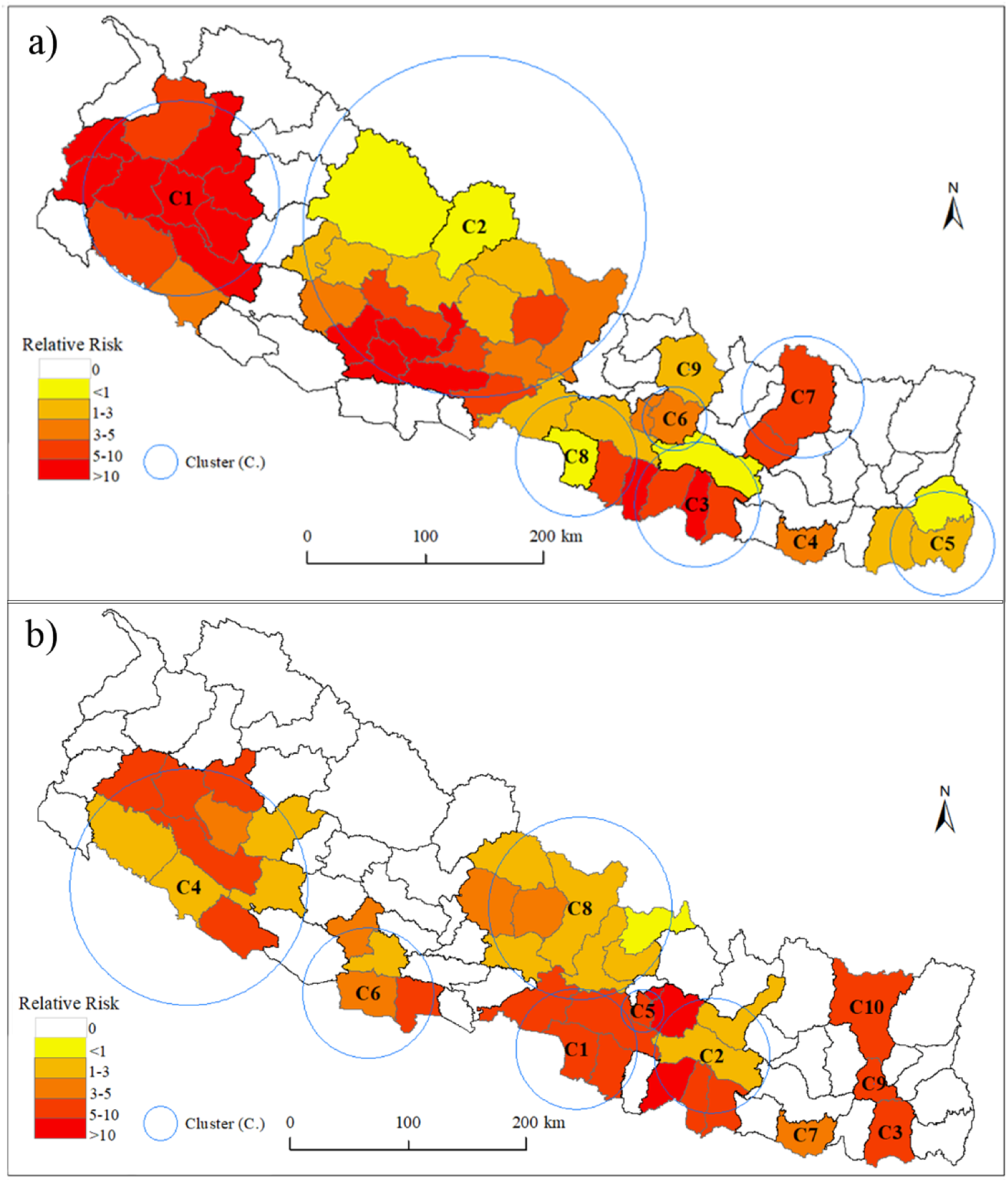
Spatial distribution of emerging space-time clusters of COVID-19 at District level. a) From 23 January – 21 July 2020; and b) 23 January – 31 August 2020.

### 3.3 Emerging district level clusters: 23 January-31 August 2020

Ten statistically significant emerging space-time clusters were detected between 23 January and 31 August 2020. Table 2 summarizes the characteristics of these cluster in terms of size, onset time, duration and relative risk level. Clusters 4, 7, 8 emerged from July 21 and persisted till the end of the study period while cluster 9 and 10 were emerged lately and persisted only for few days. The cluster 1, which is the most likely cluster, emerged on June 30 while clusters 2 and 3 arose on first week of August. Relative risk also varied significantly among these clusters. Cluster 5 (RR =10.5) had the highest relative risk followed by cluster 3, 2, and 1; while 4, 6, 7, and 9 were the clusters with lower relative risk.

**Table 2.** District level emerging space-time clusters of COVID-19 from 23 January to 31 August 2020 in Nepal (RR: relative risk). All results are statistically significant at P<0.001.

| Cluster | Radius | Start Date | End Date | # Districts | Observed | Expected | RR |
| --- | --- | --- | --- | --- | --- | --- | --- |
| 1 | 51.33 | 2020/7/30 | 2020/8/31 | 4 | 3815 | 569.21 | 7.31 |
| 2 | 49.00 | 2020/8/8 | 2020/8/31 | 6 | 2469 | 320.65 | 8.15 |
| 3 | 0.00 | 2020/8/4 | 2020/8/31 | 1 | 1280 | 133.03 | 9.91 |
| 4 | 100.35 | 2020/7/21 | 2020/8/31 | 10 | 2829 | 742.70 | 4.03 |
| 5 | 18.13 | 2020/8/11 | 2020/8/31 | 2 | 971 | 94.53 | 10.51 |
| 6 | 55.39 | 2020/8/9 | 2020/8/31 | 4 | 1066 | 232.68 | 4.68 |
| 7 | 0.00 | 2020/7/21 | 2020/8/31 | 1 | 430 | 129.15 | 3.36 |
| 8 | 77.67 | 2020/7/21 | 2020/8/31 | 8 | 752 | 345.89 | 2.20 |
| 9 | 0.00 | 2020/8/29 | 2020/8/31 | 1 | 19 | 2.10 | 9.07 |
| 10 | 0.00 | 2020/8/27 | 2020/8/31 | 1 | 20 | 3.44 | 5.82 |

Fig. 4b illustrates extent and spatial distribution 10 emerging space-time clusters of COVID-19 at the district level in Nepal between January 23 and August 31, 2020. Cluster 4 is the largest cluster with 100 km radius, which contains 10 districts of Karnali and Sudoorpaschim province. The most likely cluster (Cluster 1) was located in central Nepal covering Parsa, Makawnpur Bara and Chitwan districts. Cluster 3, 7, 9 and 10 were single districts clusters located in the Eastern Nepal. Figure 5 also elucidates the elevated risk of 38 districts lying inside these clusters with varying risk level ranging from 2.2 (Cluster 8) to 10.51 (Cluster 5). Bhaktapur, Lalitpur and Sarlahi were the districts with higher relative risk (RR> 10). The number of the districts with moderate relative risk were 16 (RR= 5-10) while lower risk (RR= 1-5) were observed 19 districts. Other 39 districts exhibited no elevated risk of exposure (RR = 0) to the COVID-19 infection.

**Figure 5.**
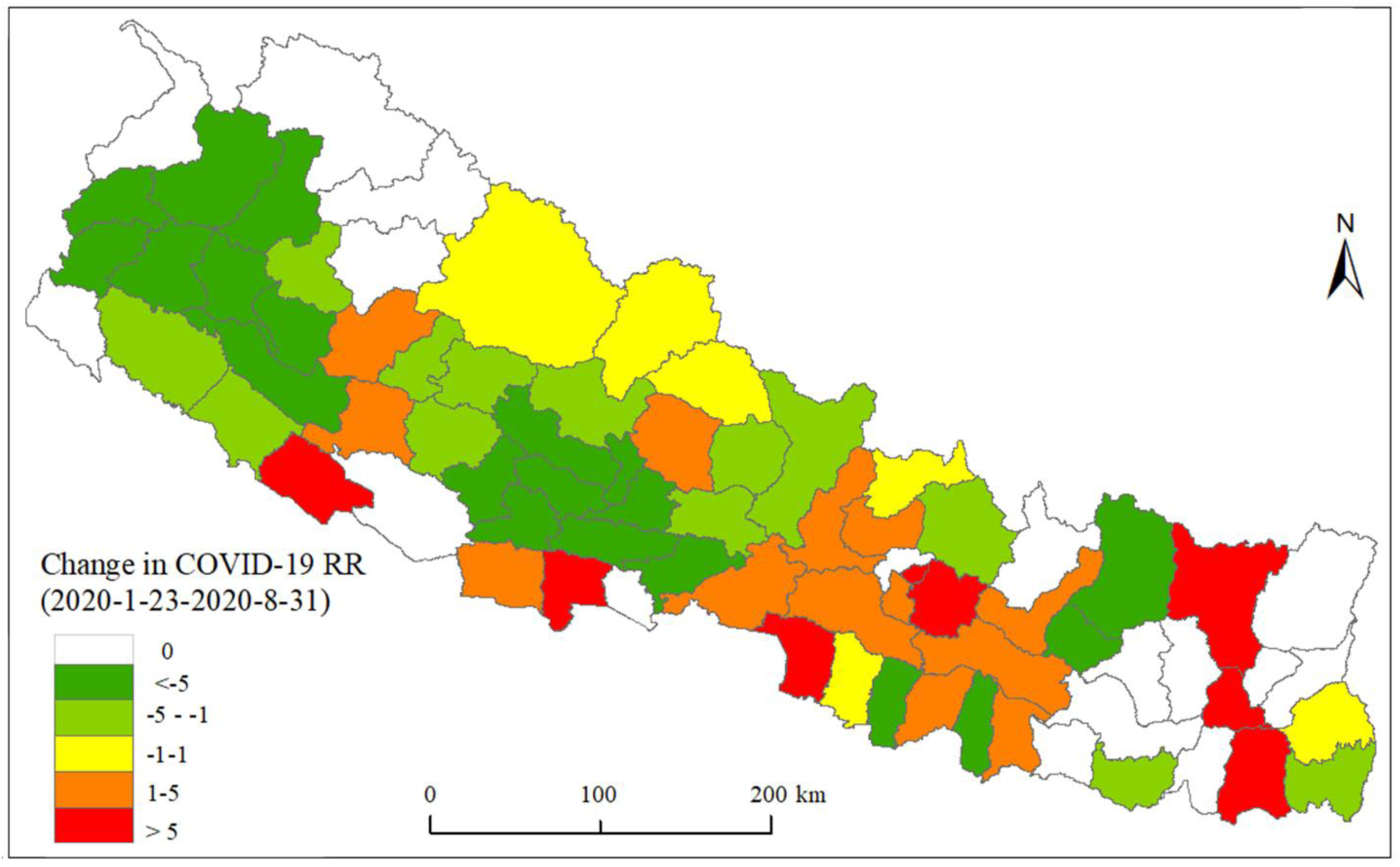
Changes in relative risk (RR) of COVID-19 between two emerging periods 23 January – 21 July and 23 January – 31 August 2020 in Nepal

### 3.4 Progression of relative risk of COVID-19 in Nepal

The changing patterns of relative risk (RR) over two emerging periods have been shown in the Figure 5. An abrupt reduction of RR was observed in 19 districts of which most of the districts belonged to the cluster 1 and cluster 2 during 23 January-21 July. A rapid rise of RR (>5) also noticed in 8 districts including Dailekh, Arghakhanchi, Baitadi, Dadeldhura, Rautahat, Parbat, Gulmi and Mahottari while moderate rise and fall in RR was observed in 11 and 12 districts symbolized by light red and light green, respectively. Some districts with RR = 0 over the two periods indicated no difference in relative risk which were regarded as “non-emerging” COVID-19 districts. However, it should be noted that these districts had also experienced the outbreak during the study period. Some of them became emerging clusters (with elevated RR) at some point in time when scanned over a shorter temporal window (Figure 6).

**Figure 6.**
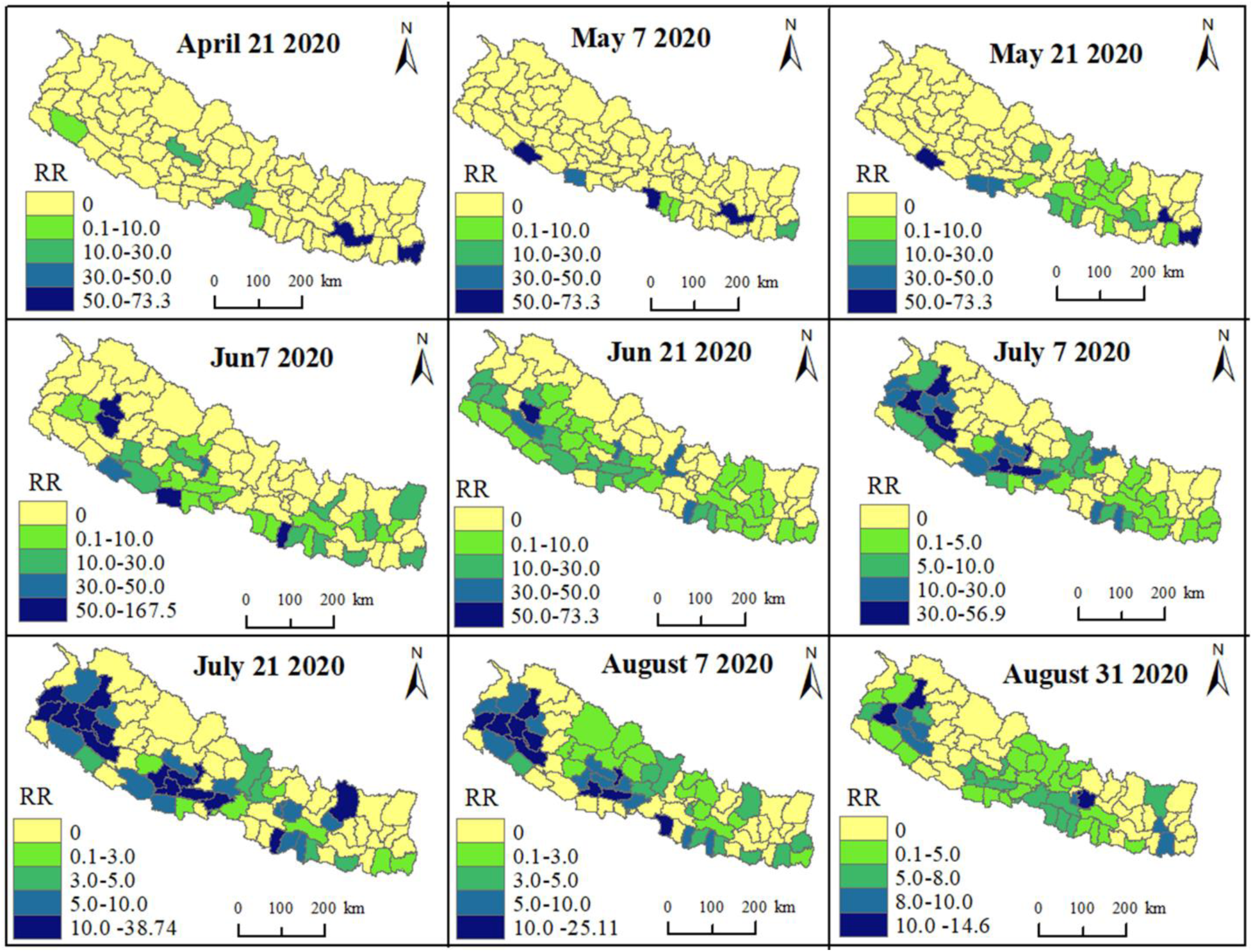
Space-time propagation of COVID-19 relative risk in different weeks in Nepal

Biweekly spatiotemporal variations of COVID-19 transmission at district level in Nepal from January 23-August 31, 2020 have been shown in the Figure 6. This short temporal window scanning enabled us to assess space-time progression of COVID-19 by locating dispersing risk and newly emerged high-risk areas along the timeline.

The elevated risk of COVID-19 transmission was observed in 21 April for the first time in Nepal although sporadic cases were reported from different districts since its first report in 23 January 2020. In this time, only six districts had elevated risk with significant variation in RR. The RR of Udayapur was extremely high (RR=297) followed by Jhapa (RR=68). The relative risk of Baglung, Chitawan, Kailali, Parsa and Nawalparasi East were moderately low (RR<20). This was the first cluster level transmission (14 cases) of COVID-19 suspected in Udayapur district of Nepal with migration history of the infected people from India. Two weeks later Banke and Parsa districts bordering India became hotspots while the risk of transmission in Udaypur persisted continuously (RR=86). By the May 21, the elevated risk expanded in 21 districts with significant spatial variation on RR. Dhankuta and Jhapa emerged as new hotspots while elevated risk of Banke remained constantly high (RR= 100). The COVID-19 transmission further spread in the next two weeks, till 07 June and the number of elevated risk (RR>1) districts reached 32. At this time Kalikot and Dailekh districts became hotspots (RR>150) while the risk of Rautahat and Kapilvastu was also significantly high (RR>50). In the later weeks, elevated risk of COVID-19 further expanded but the unexpectedly high RR was more stabilize. From the beginning of July, the districts with higher relative risk further expanded on the proximity of Dailekh and Kailkot and the vicinity of Palpa and Syangja which continue until the first week of August. By the August 31, which is the last date of study period, the elevated RR was expanded to 56 districts.

## 4. Discussion

COVID-19 pandemic has greatly affected the south-Asian countries including Nepal where number of cases are still (first week of October) increasing exponentially (MoHP, 2020). This study employed prospective space-time scan statistics for identifying currently active or emerging clusters of COVID-19 at the district level in Nepal, providing results at two distinct time periods of differential intervention attempts. The prospective scanning is valuable surveillance tool in monitoring disease outbreaks and locate active elevated risk of disease in space and time (Desjardins et al., 2020). Our findings can be useful for rapidly monitoring evolving space-time patterns of COVID-19 that will enable government and health officials to take appropriate time-sensitive intervention by considering disease’s space-time diffusion pathways and potentially prepare for future outbreaks of a highly contagious disease (Masrur et al., 2020).

After the first recorded positive case of COVID-19 on 21 January, there was a lag period of two months for the very next case. However, the cases increased after the third week of the March and continued to grow exponentially. The first ascend on the epidemic curve was observed after the third week of April when Nepalese migrants working in India returned home after nationwide lockdown there. The open border between the countries and surge of large number of returnees made it impossible to regulate the movement and manage proper test and isolation. Therefore, most of the cases were recorded from the districts of Province No 2 bordering India and that of Sudoorpaschim province, large number of people from those districts are working in India since long (Chalise 2020). Human mobility and control strategy determine the spatial spread of the epidemics (Arimura et al., 2020; Drake et al., 2020; Kraemer et al., 2020; Rader et al., 2020; Zhou et al., 2020). The areas close to the outbreak has a higher risk of contagion, especially in the initial stage of infection (Carteni et al., 2020). Indian cities were severely affected of the COVID-19 since early April (Ray et al., 2020; Tomar and Gupta 2020) and inflow of infected but asymptomatic people from those areas without testing increased the cases in particular areas of Nepal. Additionally, those districts of elevated incidences are also characterized by higher population density, lower literacy rates, higher poverty, and in turn preeminent vulnerability to the epidemics (Khanal et al., 2020). Population density is one of the important factors in shaping the spatial pattern of the epidemics as the crowded cities worldwide could experience more prolonged epidemics (Rader et al., 2020). Similar results were observed in China where the population inflow from Wuhan, the epicenter of the pandemic and the strength of economic connection were the main factors affecting the epidemic spread (Xie et al., 2020).

The spatial analysis and predictive modelling of the evolution of the COVID-19 is important to interpret the epidemic phenomenon (Franch-Pardo et al., 2020). Our prospective space–time scanning analysis revealed nine major emerging clusters for the first phase of the study (23 January-21 July). The most likely cluster, the C1, emerged on 12 June that included 11 districts from Karnali and Sudoorpaschim provinces. These districts have higher poverty and majority of the households have one or more members of the family working as low-skilled manpower in Indian cities like Mumbai, Delhi, and others (Khanal et al., 2020). The first cluster of COVID-19 observed was the consequence of infected returnees from India. Despite the nationwide lockdown imposed and borders sealed, people from India used resumed Indian railway services after the middle of May and returned back Nepal crossing the open border without taking proper precautions and in many cases violating isolation and quarantine protocols of federal and local governments (Chalise, 2020). Clusters 1-5 began on the second week of June and persisted till 21 July that were all associated to the inflow of people from India. It has signified the importance of social distancing and movement restrictions in containing the epidemic.

The prospective space–time scanning analyses for wider temporal scale, i.e. from 23 January to 31 August revealed 10 emerging clusters. The Cluster 4 in the far-western lowland Nepal was the largest cluster encompassing 10 districts with relative risk of 4.03, which is apparently the continuation of the Cluster 1 of the previous time frame i.e. 23 January-21 July 2020. The Cluster 5 with the highest relative risk of 10.5 included two districts of the capital city Kathmandu valley-Bhaktapur and Lalitpur. The number of COVID-19 cases were much higher in the Kathmandu district of the valley (MoHP, 2020); however, our analysis didn’t account a high relative risk for it due to an enormous base population. When the Government of Nepal lifted the nationwide lockdown on 21 July, people from different districts rushed to the capital city and the number of cases raised abruptly that developed a strong cluster (Cluster 5) with very high relative risk. Epidemics in crowded cities disperse rapidly and have larger total attack rates than less populated cities (Rader et al., 2020). To better understand the COVID-19 transmission dynamics, datasets on patient’s travel and contact history need to be incorporated (Masrur et al., 2020), however, there is no proper mechanism of tracking in Nepal. Therefore, the Kathmandu valley with more than four million population within 665 km^2^ area is under severe risk of COVID-19 outbreak.

During nationwide lockdown imposed by the federal government of Nepal, all public places remained shut down and strictly followed government directives. Many local municipal governments also efficiently implemented the closure, isolation, tracking and quarantine; those which failed to do so experienced initial community outbreaks. Therefore, till June 2020, community level transmission was localized in few districts such as Udaypur, Parsa, and Banke (Figure 6). However, since July, many districts of Lumbini and Sudoorpaschim provinces experienced a high relative risk. Major reason behind such was unpreparedness of the federal government (Thakur et al., 2020) which failed to seal the southern border that imported hundreds of COVID-19 positive people from India, and could not properly test, track and isolate the individuals rescued from the Arabian countries. Another important shortcoming was the use of less reliable and inefficient antibody-based diagnosis (the rapid diagnostic tools) (Bisoffi et al., 2020; Ghaffari et al., 2020) emphasized in place of the antigen-based RT-PCR. By the end of August, densely populated Kathmandu valley that had very few cases of COVID-19 for the first five months of lockdown started having thousands of cases diagnosed every day. Centralization of the health facilities in the capital city Kathmandu caused people to move into it for the diagnosis and treatment of diseases including COVID-19. It is an established fact that people having compromised immunity due to pulmonary and cardiovascular disorders are highly prone to the COVID-19 infection (Fang et al., 2020; Zheng et al., 2020). Large number of old-age people visiting hospital for medical checkup were found positive to the COVID-19 and many were diagnosed positive only after death. Inefficient and inadequate intervention against the epidemic has resulted a strong cluster within the Kathmandu valley and Bharatpur where medical facilities are centralized but becoming short to contain the COVID-19. Therefore, together with medical care, non-pharmaceutical interventions such as travel restrictions, tracking and isolation are inevitable.

## 5. Conclusion

The epidemic spread rate in Nepal has an evident spatial variation. Districts of Sudoorpaschim province and Province No 2 bordering India experienced rapid transmission of the COVID-19 when the Nepalese migrants returned home in May/June. The unmanaged population inflow from India crossing the sealed border had significant effects on the epidemic spread rate. The capital city Kathmandu and Bharatpur where medical facilities are concentrated have become the highest-risk active clusters since August. It is important to detect emerging clusters that would reveal more updated space-time transmission dynamics of COVID-19 to better allocate resources and improve decision-making as the outbreaks continue to grow. The purposive and time-bound movement restriction appears to be the most important non-pharmaceutical intervention against the COVID-19 for resource-scarce countries with limited health care facilities.

## Data Availability

All data used in the manuscript are freely available.

## Acknowledgement

We thank Ministry of Health and Population, Government of Nepal for providing the valuable data on COVID-19 in Nepal.

## Author Contributions

BKA, LK and HL conceptualized the study. BKA, LK, SKA, SP, BKN and BKP collected and processed data. BKA and LK analyzed the data and prepared the manuscript. ASMM, ZR and YY helped in manuscript improvement. HL supervised the overall study and provided multiple revisions. All authors read and approved the final manuscript.

## Conflicts of Interest

The authors declare no conflict of interest.

## Notes

### Competing Interest Statement

The authors have declared no competing interest.

### Funding Statement

National Natural Science Foundation of China (82041021).

